# Autism Caregiver Coaching in Africa (ACACIA): Protocol for a type 1-hybrid effectiveness-implementation trial

**DOI:** 10.1101/2023.09.10.23295331

**Authors:** Lauren Franz, Marisa Viljoen, Sandy Askew, Musaddiqah Brown, Geraldine Dawson, J Matias Di Martino, Guillermo Sapiro, Katlego Sebolai, Noleen Seris, Nokuthula Shabalala, Aubyn Stahmer, Elizabeth L Turner, Petrus J de Vries

## Abstract

**Background:** While early autism intervention can significantly improve outcomes, gaps in implementation exist globally. These gaps are clearest in Africa, where forty percent of the world’s children will live by 2050. Task-sharing early intervention to non-specialists is a key implementation strategy, given the lack of specialists in Africa. Naturalistic Developmental Behavioral Interventions (NDBI) are a class of early autism intervention that can be delivered by caregivers. As a foundational step to address the early autism intervention gap, we adapted a non-specialist delivered caregiver coaching NDBI for the South African context, and pre-piloted this cascaded task-sharing approach in an existing system of care.

**Objectives:** First, we will test the effectiveness of the caregiver coaching NDBI compared to usual care. Second, we will describe coaching implementation factors within the Western Cape Department of Education in South Africa.

**Methods:** This is a type 1 effectiveness-implementation hybrid design; assessor-blinded, group randomized controlled trial. Participants include 150 autistic children (18-72 months) and their caregivers who live in Cape Town, South Africa, and those involved in intervention implementation. Early Childhood Development practitioners, employed by the Department of Education, will deliver 12, one hour, coaching sessions to the intervention group. The control group will receive usual care. Distal co-primary outcomes include the Communication Domain Standard Score (Vineland Adaptive Behavior Scales, Third Edition) and the Language and Communication Developmental Quotient (Griffiths Scales of Child Development, Third Edition). Proximal secondary outcome include caregiver strategies measured by the sum of five items from the Joint Engagement Rating Inventory. We will describe key implementation determinants.

**Results:** Participant enrolment started in April 2023. Estimated primary completion date is March 2027.

**Conclusion:** The ACACIA trial will determine whether a cascaded task-sharing intervention delivered in an educational setting leads to meaningful improvements in communication abilities of autistic children, and identify implementation barriers and facilitators.

**Trial registration:** NCT05551728 in Clinical Trial Registry (https://clinicaltrials.gov)

## Introduction

Globally there is growing recognition of the importance of strengthening the capacity of caregivers and clinicians to deliver services at the community-level that result in improved quality of life of all autistic people [1]. Early autism intervention is recognized as critical given that specialist-delivered intervention can significantly improve child and family outcomes, reduce the need for services at a later age, and be a cost-effective approach [2–3]. Most autistic people live in low- and middle-income countries (LMIC), where few systems of care provide developmental screening or early intervention [4]. Forty percent of the world’s children will live in Africa by 2050 [5]. This demographic reality underlines the necessity of feasible early intervention services that can be scaled-up within existing systems of care.

Research has demonstrated that caregivers can be coached to implement early intervention strategies with their young autistic child across daily activities [6]. Low intensity caregiver coaching can impact caregiver-child interaction styles and child outcomes [7–8]. Naturalistic Developmental Behavioral Interventions (NDBI) comprise a class of early intervention approaches that caregivers can deliver [9]. The Early Start Denver Model (ESDM) is an evidence-based NDBI [10]. In the ESDM approach, strategies increase a child’s attention and engagement with their caregiver, thereby increasing opportunities for learning communication and social skills, leading to improvements in child outcomes, including language abilities [11].

The efficacy of ESDM delivered via therapists, caregivers, and teachers has been evaluated in a wide range of cultures, including in the United States, Israel, Taiwan, Italy, China, Japan, and Australia among others [2, 10, 12–20]. While these studies indicate that NDBI approaches, such as ESDM, hold promise for addressing the needs of young autistic children in LMIC, the research outlined in the current protocol paper aims to address three important gaps in the existing literature. First, very few studies to date have included diverse community partners, including autistic self-advocates, in the research process [21]. Such involvement is important to ensure that the intervention is acceptable to the autism community [22]. Second, caregiver coaches have almost exclusively been highly trained specialists, usually licensed in the fields of psychology and/or education. Lack of access to such specialists in most parts of the world requires that studies include task-sharing to non-specialists as an implementation strategy [1, 23].

Third, in a meta-analysis of early autism interventions using non-specialist, only two of the included studies were from LMIC [24–26]. Furthermore, in these two studies the therapists and teachers were the identified ‘non-specialists’, a workforce unlikely to be scalable in Africa. In an Indian study, supervised lay health workers delivered a developmental intervention, demonstrating positive effects on children’s outcomes [27]. Although that study supports the feasibility of non-specialist coaches in India, barriers and facilitators of implementation will be different in other world regions, including Africa. Thus, we aim to develop an early autism intervention approach in Africa that is inclusive of diverse perspectives and incorporates cascaded task-sharing.

In Africa, Community Health Workers are often proposed as the non-specialists to target for task-sharing [28]. In some LMIC, early childhood development services have been integrated into maternal and child Community Health Worker programs [29–30]. Challenges with this approach have been identified and include limited Community Health Worker training in early child development, difficulties managing competing demands resulting in poor quality of care and burn out, and lack of individualization of approach [31–33]. This lack of individualization is particularly problematic for children with variable developmental profiles, a common observation in autism. In the current study, we aim to address each of these challenges by working with Early Childhood Development (ECD) practitioners, a non-specialist workforce distinct from Community Health Workers, who have specific training in early child development. ECD practitioners are employed by the Education Department, will have dedicated time to coach, will work under supervision, and use an intervention that can be tailored to meet individual needs. Addressing these challenges will help create a non-specialist workforce, well-positioned to provide coaching for caregivers to deliver early autism intervention, thereby increasing the likelihood of long-term sustainability and scalability of the approach.

South Africa, with one of the highest income inequality rates worldwide, is a diverse upper-middle income country [34]. Few families with young children are able to access early autism intervention due to limited services and structural barriers, including poverty and the legacy of apartheid [35–40]. Families able to access publically available services receive approximately one 30-minute therapy session every 4–6 weeks for a limited period of time. Caregivers are typically not included in therapy sessions and are asked to wait outside the room. A South African study documented racial variations in expressive language in a neurodevelopmental clinic, with a significantly greater number of Black compared to White children non-verbal at diagnosis (94% vs 42%) [41]. The need for early autism intervention is increasing, with a documented 276% rise over the last decade in the children waiting for special education services [37].

As a foundational step to address the intervention gap in South Africa, in 2015 a partnership between Duke University in the United States and the University of Cape Town (UCT) in South Africa was established [42]. The long-term goal of this partnership was to contextually adapt and implement a cascaded task-sharing NDBI for young autistic children, and to integrate it into a South African system of care. The research team completed the following steps: First, after a significant body of formative research, ECD practitioners were identified as the non-specialist workforce that could serve as intervention coaches [36]. ECD practitioners are well-suited for this role as they are employed by the Education Department, trained in early child development, supervised in their day to day activities, and supported by national policy [43]. A partnership between the ECD workforce and the National Education Department was already established – an alignment that will be critical for future scale-up [44]. Second, the specific NDBI approach (ESDM) was identified based on local interest and available resources. Specifically, this approach was selected because UCT-based clinicians had received some training in ESDM when the partnership with Duke University was established, and the US-based Principal Investigator (PI) was a faculty member at an academic institution where ESDM training occurred. Furthermore, the relationship-based approach of the ESDM aligns with the dominant approach to early childhood mental health in South Africa [45]. Third, the ‘fit’ of NDBI-caregiver coaching with South African caregiver-child dyads was evaluated [35, 46–48]. Interviews with caregivers and behavioral coding of caregiver-child interactions were conducted to understand interaction styles, and common daily routines in which the caregiver coaching intervention could be carried out. Fourth, ESDM coaching procedures were adapted for delivery by non-specialists. Fifth, the team conducted a pilot study of the cascaded task-sharing approach was completed with 10 caregiver-child dyads [23]. Cascaded task-sharing refers to task-sharing the caregiver-coaching role, from a highly trained specialist to a non-specialist. This pilot study demonstrated that caregivers could learn NDBI strategies when coached by non-specialist ECD practitioners, with caregiver implementation fidelity increasing in 10/10 participants. Growth in child communication was also documented, with significant gains seen on the Communication Domain Standard Score of the Vineland Adaptive Behavior Scales, Third Edition (VABS-3) [49] (9/10 improved), and the Language and Communication Developmental Quotient of the Griffiths Scales of Child Development, Third Edition (Griffiths III) [50] (9/10 improved). Finally, mindful of the ‘digital divide’, intervention materials were adapted to be shared through WhatsApp, a low-cost messaging software widely used across Africa.

Given that the contextually adapted cascaded task-sharing NDBI has demonstrated proof-of-principle, a larger scale trial is required to answer a set of key questions. The first key question is whether the caregiver coaching intervention is effective in terms of its ability to improve child outcomes meaningfully. The majority of caregiver coaching intervention studies only utilize study-specific behavioral coding of caregiver-child interaction as a primary outcome measure [51]. This approach has significant limitations. Behavioral coding measures may only identify circumscribed and temporary changes that may not be indicative of broader developmental gains. A recommended approach to assess meaningful intervention impact is to use tools that measure change that is clinically meaningful [52]. This can be accomplished by using child-focused measures that identify generalization of child skills with different interaction partners and environments (i.e., clinician-administered standardized developmental assessments). The second key question to address is identification of intervention implementation facilitators and barriers. The implementation context is critical to consider in early autism intervention, particularly the ‘fit’ between intervention and context [35]. The Exploration, Preparation, Implementation, Sustainment (EPIS) framework, is an implementation framework that has been used to specify the complex context for evidence-based practice implementation [53, 54]. The EPIS framework can guide identification of influential contextual factors that hinder or support the implementation process. Recognizing the importance of community partner engagement, the EPIS framework includes a community academic partnership as a bridging factor that can address potential ideological differences, help ensure research products are acceptable to end-users, and support longitudinal community partner engagement across the research cycle [54, 55].

For the current study, we will conduct a 1:1 parallel-arm, single (assessor)-blinded, individually randomized, group-treatment, randomized controlled trial (RCT) using a hybrid type 1 effectiveness-implementation design [56, 57]. We will evaluate the effectiveness of the cascaded task-sharing intervention on child communication outcomes compared to usual care, in young autistic children (18-72 months) who live in Cape Town, South Africa, while documenting factors that affect implementation. ECD practitioners will complete a four-day training with ESDM-certified therapists from South Africa. During the training ECD practitioners will be introduced to core NDBI concepts and caregiver coaching strategies. ECD practitioners will practice these new coaching and NDBI strategies with a caregiver-child dyad. Ongoing supervision is follows the apprenticeship model for lay counsellor supervision in mental health [58]. The RCT design can be classified as an individually randomized group-treatment trial in which ‘groups’ arise in the treatment arm as a result of multiple caregivers being coached by the same ECD practitioner (with coaching provided individually to the caregiver) [57]. This can potentially result in correlated outcomes of caregiver-child dyads in the intervention arm, trained by the same ECD practitioner (i.e. these caregivers would therefore be members of the same ‘group’). We note that no ‘grouping’ and therefore no correlation of outcomes are expected in the control arm as no training or interactions with the same ECD practitioner are expected. The primary hypothesis of the proposed study is that young autistic children who are randomized with their caregiver to receive 12, 1-hour caregiver coaching sessions (intervention arm), will have greater gains in communication abilities (assessed using two co-primary outcomes, namely the Communication Domain Standard Score from the VABS-3 and the Language and Communication Developmental Quotient from the Griffiths III), at distal follow-up assessment (24 weeks post-randomization), compared with autistic children who receive usual care (control arm). The secondary hypothesis of this study is that caregivers in the intervention arm will have greater gain in NDBI caregiver strategies (measured by the sum of 5 caregiver items from the Joint Engagement Rating Inventory during caregiver-child interaction) [59], at a proximal follow-up assessment (16 weeks post-randomization), compared to the control arm.

## Materials and methods

See S1 Checklist for the Standard Protocol Items Recommendations for Interventional Trials (SPIRIT) Checklist used for study protocol reporting [60]. The schedule of enrolment, baseline assessment, intervention, and 16 weeks (proximal) and 24 weeks (distal) follow-up post-randomization assessments is shown in Fig 1.

**Fig 1.**
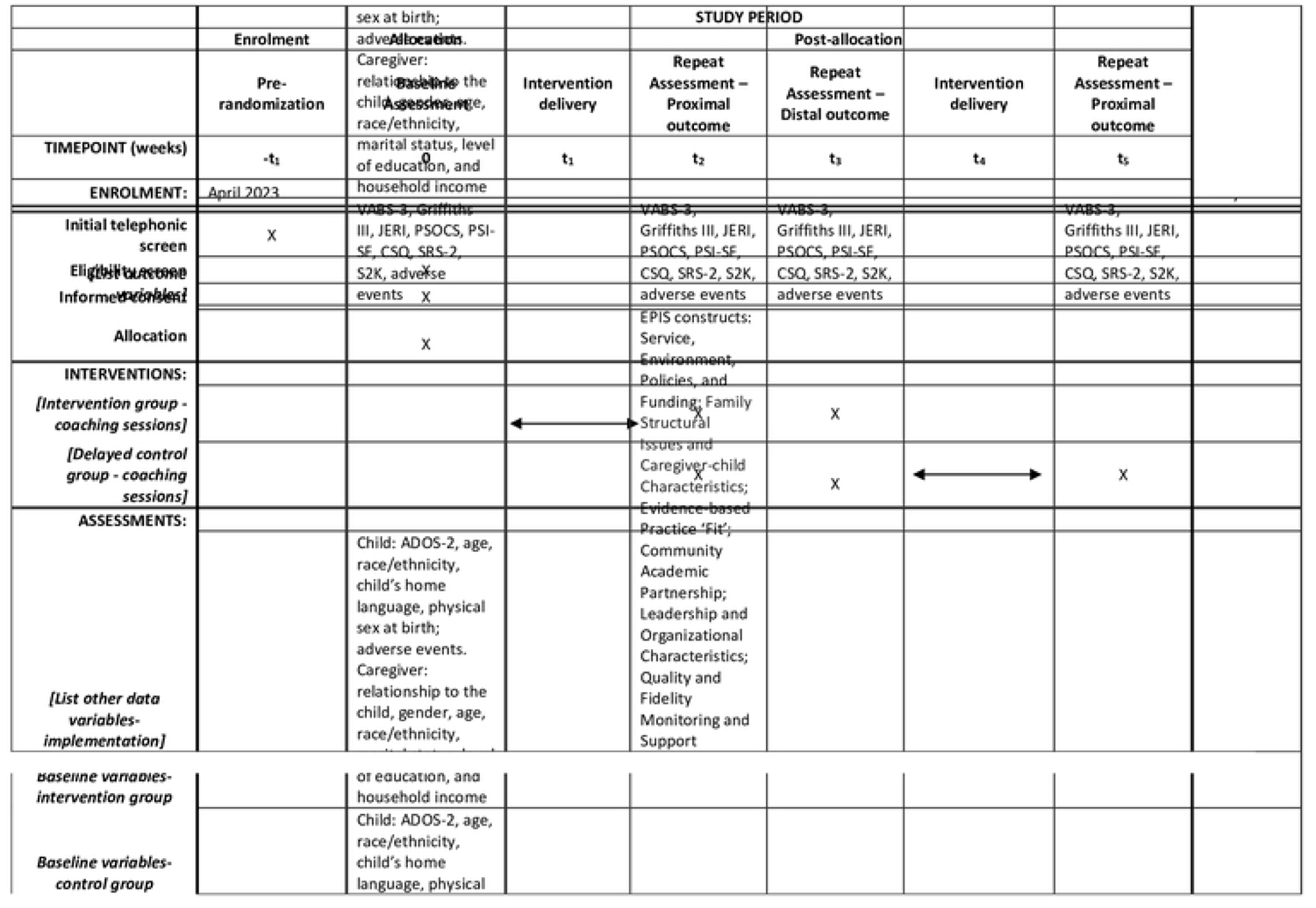
Standard Protocol Items: Recommendations for Interventional Trials (SPIRIT) schedule of enrolment, interventions, and assessments (ADOS-2: Autism Diagnostic Observation Schedule, Second Edition; VABS-3: Vineland Adaptive Behavior Scales-3; Griffiths III: Griffiths Scales of Child Development, Third Edition; JERI: Joint Engagement Rating Inventory; PSOCS: Parent Sense of Competence Scale; PSI-SF: Parenting Stress Index-Short Form; CSQ - Caregiver Strain Questionnaire; SRS-2: Social Responsiveness Scale, Second Edition; S2K: SenseToKnow mobile app; *-t_1_:* Pre-randomization; *0:* Baseline Assessment; *t_1_:* Intervention delivery; *t_2_:* Repeat Assessment – Proximal outcome; *t_3_:* Repeat Assessment – Distal outcome; *t_4_:* Repeat Assessment – Proximal; *t_x_:* close out)

### Trial setting

The study will be conducted in Cape Town, South Africa. Formative work identified the Education Department as study implementation partner [36]. With the support of Inclusive and Specialized Education Department Leadership, we will identify Special Education Schools in the Cape Town Metropolitan area who employ ECD practitioners, the non-specialist workforce that will deliver the cascaded task-sharing intervention. Caregiver coaching will occur in these schools. We will recruit participants for the RCT (caregiver-child dyads) from the Western Cape Education Department Provincial Autism Waiting List for special education services. This is a list of autistic children with a clinical diagnosis of autism from community providers awaiting public sector special education services when they reach school-going age [38].

### Participants

To assess intervention effectiveness, we will invite caregiver-child dyads who meet the following criteria to join the study: (1) child’s age is 18-72 months, (2) child meets the Diagnostic and Statistical Manual of Mental Disorders, Fifth Edition (DSM-5) criteria for autism spectrum disorder [61], informed by the Autism Diagnostic Observation Schedule, Second Edition (ADOS-2), [62] which is administered by research reliable clinicians, (3) child’s caregiver speaks isiXhosa, isiZulu, Afrikaans, or English, (4) child’s race is African or Coloured (a South African term for mixed race), (5) lives in the Cape Town recruitment area, and (6) caregiver is at least 18 years of age. Exclusion criteria are: (1) child has a genetic disorder of known etiology (for example, fragile X syndrome), (2) child has significant sensory or motor impairment that would preclude use of the play materials, (3) child has major physical abnormalities that would interfere with participation in the coaching intervention, a (4) child has a history of serious head injury and/or neurological condition, and (5) caregiver indicates they will be unable to attend assessments and 12 sessions. We will enroll 150 caregiver-child dyads in the RCT (300 participants).

To identify implementation barriers and facilitators we will invite those involved in intervention implementation through participation in the intervention trial and community academic partnership members to provide feedback. Specifically, caregivers who receive coaching, ECD practitioners who conduct coaching, ECD supervisors, and Western Cape Education Department leadership will be included. These participants will be directly involved in intervention implementation and be able to provide input on the specific outer (for example, policies and funding) and inner (for example, provider training and child characteristics) contextual, and innovation (for example, intervention complexity) factors outlined in the EPIS framework [54]. Community academic partnership members will include the PI, Co-PI, Education Department leadership, and other community partners (for example, caregivers and autistic adults). These participants will provide input on functioning of the community-academic partnership. We will enroll 20 participants to assess implementation barriers and facilitators.

### Trial intervention

The cascaded task-sharing intervention, Autism Caregiver Coaching in Africa (ACACIA), is informed by ‘Help is in Your Hands’ online materials, which include core components of ESDM that align with eleven common elements shared across all NDBI [63]–[65]. ‘Help is in Your Hands’ materials are open-access, designed for low-resource, low-literacy settings, and include narrated video and animation examples of caregivers using intervention strategies across daily routines in naturalistic home-based environments [64]. The research team systematically adapted both coaching procedures and ‘Help is in Your Hands’ intervention content to fit the South African context [23, 35, 36, 46–48]. Modifications to training, supervision, and session structures were made to facilitate intervention delivery by non-specialist providers. Adaptations to intervention materials produced simple visuals and text that can be sent to caregivers through WhatsApp, a low-cost messaging software, widely used across Africa. Adaptations to intervention materials were made collaboratively by multicultural/multilingual South African ESDM therapists, the PI (an ESDM therapist and certified trainer), and a co-developer of ‘Help is in Your Hands’. The Ecological Validity Framework, which consists of eight dimensions of intervention important to consider during adaptation, guided the adaptation approach [66]. Twelve caregiver-facing WhatsApp messages containing visuals with simple-text, and ECD practitioner-facing session scripts were developed. Over 12, one-hour ECD practitioner delivered coaching sessions, caregivers review ACACIA intervention content with ECD practitioners and are coached to increase their child’s attention and communication, create and vary joint activity routines, and use the antecedents, behaviors, and consequences to support the development of new behaviors. See Table 1 for a description of the specific content of the 12 ACACIA coaching sessions. Participants randomized to the control arm will receive usual care, which may vary between participants but will likely include low-frequency of speech and/or occupational therapy (i.e., approximately one 30-minute therapy session every 4–6 weeks).

**Table 1.** ACACIA coaching session content.

| ACACIA session | Session content |
| --- | --- |
| 1 | Set goals, and identify child likes and dislikes |
| 2 | Identify child spotlight of attention and how to increase attention to caregiver |
| 3 | Identify and follow child-preferred interests |
| 4 | Increase child use and understanding of non-verbal communication |
| 5 | Support child ability to share interests through giving, pointing and showing |
| 6 | Increase child use and understanding of verbal communication |
| 7 | The 4-part joint activity structure (set-up, theme, variation, and closing) |
| 8 | Sensory social routines to increase child attention and learning |
| 9 | Support child learning across all daily activities |
| 10 | Understand reasons for and consequences of child behaviour |
| 11 | Teach a new behaviour by showing, telling and helping your child |
| 12 | To help new behaviours stick, practice, get others to practice, and give less help over time |

### Measures

#### Sociodemographic and child characteristics

At baseline assessment descriptive characteristics for the child (age, race/ethnicity, child’s home language, physical sex at birth) and caregiver (relationship to the child, gender, age, race/ethnicity, marital status, level of education, and household income). Child autism-related behaviors will be assessed with the ADOS-2, a clinician administered, play-based, semi-structured assessment designed to elicit autism-related behaviours in a standardized way [62]. Adverse events will be assessed across study contact points using a general inquiry elicitation approach [67]. Additionally, caregivers will report on autism related-services (outside of caregiver coaching sessions) their child receives across study contact points.

### Clinical outcome measures

#### Primary clinical outcome

*The Vineland Adaptive Behavior Scales-3 (VABS-3) - Communication Domain Standard Score*. The VABS-3 is a caregiver interview, which is semi-structured and administered by a trained clinician. The VABS-3 evaluated how the child performs adaptive behaviors relative to age-based norms. The VABS-3 yields an overall composite score and standard scores across four domains including communication, socialization, daily living skills, and motor skills [49]. Subscale scores range from 20-140 with higher scores indicating greater frequency of the specific behavior.

*The Griffiths Scales of Child Development, Third Edition (Griffiths III)-Language Developmental Quotient*: The Griffiths-III is a developmental measure, administered by a trained clinicians, that evaluates children birth to six years of age [50]. The Griffiths-III profiles child development across 5 domains: language and communication, foundations of learning, personal-social-emotional, eye and hand coordination and gross motor. Developmental quotients (DQ) are calculated by (Developmental Age/Chronological Age) *100. Higher DQ indicate greater consistency between chronological and developmental age.

#### Secondary clinical outcomes

*The Joint Engagement Rating Inventory (JERI) - Sum of five Caregiver Activity Items*: is a behavioral coding inventory that assesses specific features of caregiver-child interactions [59]. The JERI includes 32, 7-point Likert scale items, which assess dyadic interaction, child activity, child engagement states and caregiver activity. In the ACACIA trial, a composite score of five JERI caregiver activity items will be calculated and include: following the child’s interests, scaffolding, language facilitation, affect, and communicative temptations. For each of the 7-point caregiver activity items, higher scores will indicate higher frequency and quality of caregiver behaviors.

#### Additional measures

*The Parent Sense of Competence Scale (PSOC)*: is a 16-item questionnaire, completed by the caregiver, that measures feeling of confidence and competence in ‘parenting’ their children and ‘parenting’ satisfaction [68]. Items, across these two domains, are rated on a 6-point Likert scale with high scores representing high degrees of satisfaction and efficacy.

*The Parenting Stress Index-Short Form (PSI-SF)*: is a 36-item questionnaire, completed by the caregiver, and designed to measure stress associated with parenting [69]. Three subscales (difficult child, parent–child dysfunctional interaction, and parenting distress), with 12 items per subscale, are rated on a 5-point Likert scale, with higher scores indicating a greater degree of parenting stress.

*The Caregiver Strain Questionnaire (CSQ)*: is a 21-item, 5-point Likert scale caregiver-completed measure of strain experienced by the caregiver [70]. Higher scores relate to higher caregiver strain.

*Social Responsiveness Scale Second Edition (SRS-2)*: is a 65-item, 4-point Likert scale caregiver-completed measure of social-related challenges in autism [71]. Five subscales (social cognition, social awareness, social motivation, social communication, and restricted interests and repetitive behaviour), and a Total Score are reported, with higher scores indicating greater challenges.

*SenseToKnow*: is a digital assessment method, that contains a set of brief stimuli, displayed on a tablet, designed to elicit multiple autism-related behaviors including attention to speech, social attention, affective expression, fine motor skills and postural control [72–77]. Using recordings made by the camera in a tablet, a computer automatically codes these behaviors using computer vision analysis and machine learning.

### Implementation outcome measures

Fig 2 outlines specific EPIS framework outer and inner contextual, bridging and innovation factors, selected to evaluate based on the teams formative work (See Fig 2) [54].

**Fig 2.**
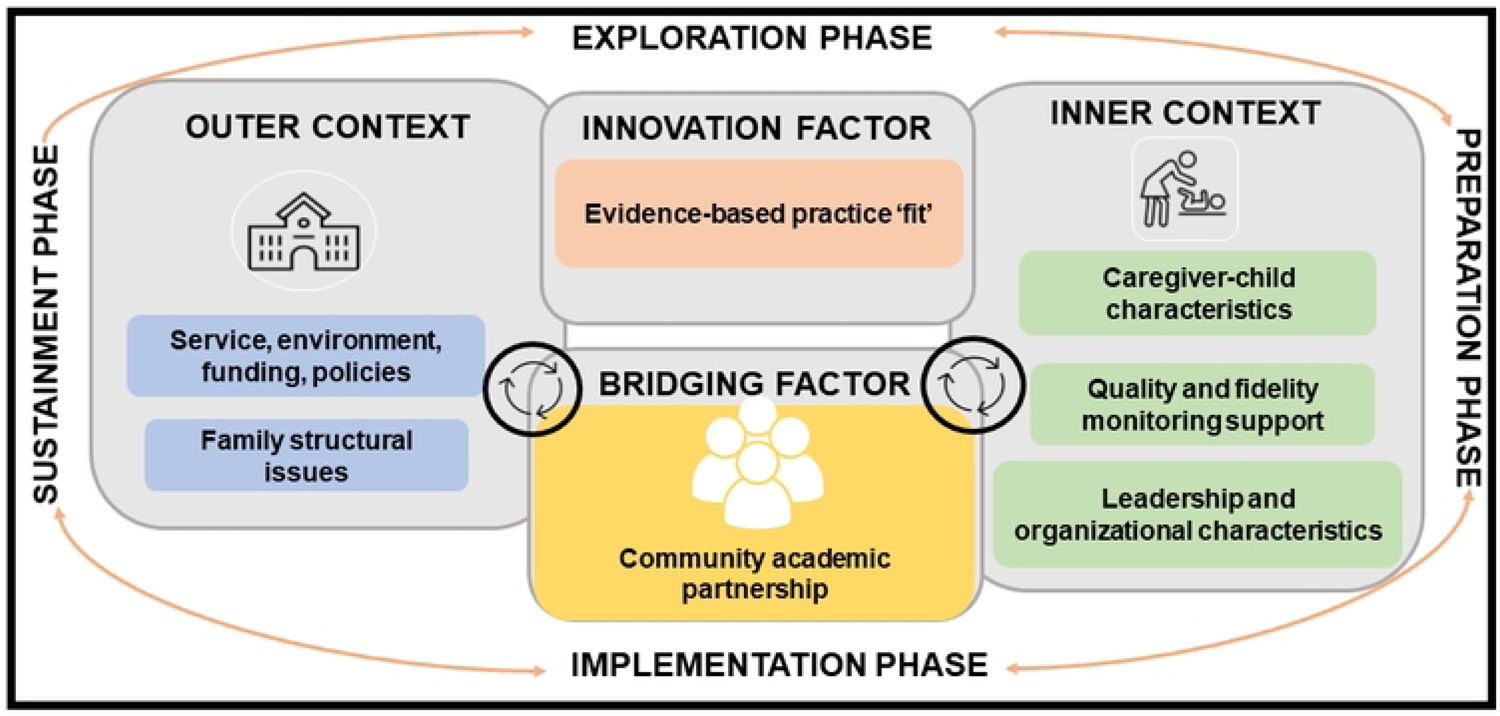
EPIS constructs to identify implementation determinants for scale-up

#### Service, environment, policies, and funding

Individual interviews and focus groups will be completed with personnel involved in intervention implementation. Qualitative questions will assess (within the Education Department): (1) the feasibility of external autism early intervention trainings, monitoring and support, (2) funding avenues that could facilitate external trainings and ongoing monitoring and support, (3) relevant policies that relate to children with developmental disabilities, and (4) policy direction in the area of early child development and care of children with developmental disabilities.

#### Family structural issues and caregiver-child characteristics

Caregivers of young autistic children will complete focus groups and the Caregiver Strain Questionnaire (described above under additional measures). Qualitative questions will assess caregiver: (1) perceptions of and goals for their child, (2) perceptions of coaching, and (3) contextual factors that affect coaching implementation (for example, language, culture, location, cost, delivery agent, family support, and available resources).

#### Evidence-based practice ‘fit’

Individual interviews and focus groups will be completed with the personnel involved in implementation of the cascaded task-sharing approach. Qualitative questions will assess: (1) logistics of caregiver coaching, including impact on workload, burden and space, (2) perceptions of caregiver coaching, including child and caregiver impact, (3) perceptions of coaching materials, including WhatsApp intervention content, and (4) sustainability of coaching practices.

#### Leadership and organizational characteristics

Two brief surveys will be completed by personnel involved in intervention implementation in order to understand leadership and the organizational characteristics of the implementation partner (Education Department). The Implementation Leadership Survey is a 12-item measure that assesses support from leadership for the implementation of evidence-based practices [78]. The Implementation Climate Scale is an 18-item measure that assesses the value an organization places on implementation of evidence-based practices [79].

#### Quality and fidelity monitoring and support

Fidelity monitoring and focus groups will be completed with personnel involved in implementation of the coaching intervention. Both caregiver and coaching implementation fidelity will be assessed using ESDM fidelity checklists [80]. The 13-item ESDM Caregiver Fidelity Checklist assesses the fidelity with which caregivers use ESDM strategies during caregiver-child interactions. Each fidelity item rates performance from one to five, with higher scores reflecting a greater fidelity. The 13-item ESDM Coaching Fidelity Checklist assesses specific coaching behaviours. This checklist includes a Likert-scale of 1 to 4, with higher scores reflecting greater fidelity. Focus groups will assess: (1) feasibility of establishing fidelity of early intervention practices, (2) ongoing available support and monitoring to support fidelity, and (3) funding to facilitate ongoing support and monitoring.

#### Community academic partnership

Community academic partnership members will complete a quantitative survey and a qualitative individual interview to understanding the partnerships functioning and impact. Survey items are informed by a community academic partnership systematic review [81, 82]. Items, will be assessed across five domains: general satisfaction, impact, trust, collaborative decision making, and meeting organization and structure, with community academic partnership members selecting whether a facilitating factor was ‘present’ or ‘not present’ during the preceding year. Qualitative individual interviews will follow survey administration to further expand upon the presence or absence of facilitating factors.

### Measuring outcomes at post intervention follow-up

#### Clinical Outcomes

Data will be collected for intervention and control arm at baseline (Baseline assessment), 16-weeks post-randomization (Repeat assessment – proximal outcome), and 24-weeks post-randomization (Repeat assessment – distal outcome). Control arm dyads will then receive the intervention. Data collection will be performed for the control arm 36-weeks post-randomization (Repeat assessment – proximal). Fig. 3 shows the RCT study design. All data collection will occur through in-person assessments by assessors blinded to participant study arm allocation.

**Fig 3.**
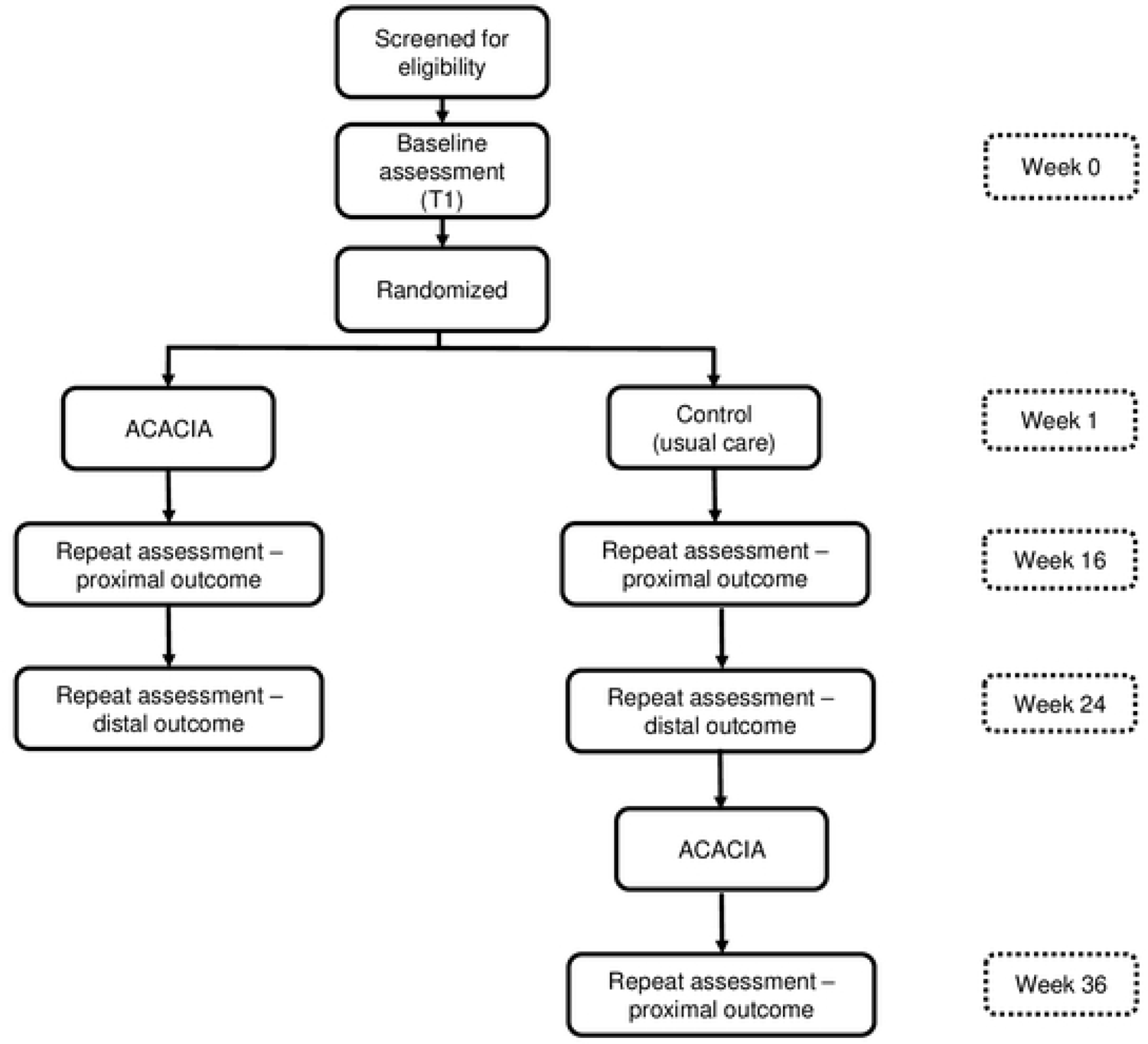
Flow diagram of participant enrolment, group allocation, and follow-up of the individually randomized group treatment RCT with delayed intervention control

#### Implementation Outcomes

To understand implementation barriers and facilitators, data will be collected from those involved in intervention implementation and community academic partnership members. Data for caregivers in the intervention group will be collected 16-weeks post-randomization (Repeat assessment – proximal outcome). Data for ECD practitioners, and other Education Department team members will be collected at a single time point (mid-year in Year 3 of the study). Data for community academic partnership members will be collected at a single time point in Years 2 and 4 of the study.

#### Sample Size

We target a minimum of 80% power for each of the two co-primary clinical outcomes used to evaluate effectiveness. More specifically, we power each co-primary clinical outcome at a two-tailed alpha level of 0.025 based on an individually randomized group-treatment trial design with 150 recruited dyads (75 in each of the two trial arms). We further assume a common outcome variance in each arm, conservatively anticipate an intraclass correlation (ICC) of 0.02 in the intervention arm, a magnitude of clustering measured in other contexts, and assume that the coaching will be delivered by 3 ECD practitioners, each of whom will deliver the cascaded task-sharing intervention to an equal number of caregivers [57]. Using a standard approach to calculate power for an individually randomized group-treatment trial, we estimate being able to detect a standardized effect size of 0.63 for each of the co-primary outcomes, based on having data from 60 participants per arm at the distal 24-week follow-up, assuming 20% attrition of the original 75 per arm [83].

#### Recruitment

Partner schools will identify caregiver-child dyads from their waitlist of young autistic children who are not enrolled in in-person instruction. School staff will share with the research coordinator the contact information of the primary caregiver of potentially eligible dyads. The research coordinator will contact potentially eligible participants via phone to provide study information, and assess interest in study participation. Those who are interested in study participation will be screened for eligibility based on study inclusion/exclusion criteria. If the caregiver-child dyad are eligible based on phone screen inclusion/exclusion criteria, they will be invited to participate in the study and a baseline assessment will be scheduled. Implementation outcome participants will be purposively sampled and include all who receive (caregivers in the intervention arm), deliver (ECD practitioners), supervise (ECD supervisors) and support (Education Department leadership) intervention delivery, and community academic partnership members.

#### Sequence generation and randomization

Randomization will occur after the baseline visit is completed using stratification by caregiver-reported child physical sex at birth. The randomization process will be managed using REDCap, a web-based software platform that is secure and designed to support research study data capture [84, 85]. A study statistician will generate allocation tables to assign participant dyads equally (1:1) to intervention and control arms. Separate allocation tables will be generated for boys and girls using permuted blocks randomly sized from 2 to 8. These allocation tables will then be uploaded to REDCap where they will function to determine the intervention allocation of a participant when the research coordinator clicks a button in the software to trigger the randomization. The REDCap software prevents staff and participants from accessing the allocation tables to predict the intervention or control arm assignment.

Participants will learn their arm assignment after completing their baseline assessment, when they will be oriented to the intervention or informed that they are on the intervention wait list and will also be scheduled to come for follow-up assessments.

### Allocation concealment

Baseline assessments will be conducted prior to randomization to ensure masking of caregiver-child dyads to allocation status, after which concealment is no longer possible.

#### Blinding

Due to the administrative logistics of the study and intervention, participants and some study staff and investigators cannot be blinded. However, clinical outcome assessors and statisticians will be blinded participant arm assignment. Participants will be asked to not disclose arm allocation during assessments. Every effort will made to minimize unnecessary exposure of the intervention arm or components. REDCap has several features to limit the visibility of the study arm to specific staff after the intervention assignment has been made which will be utilized whenever possible. Clinical assessors performing evaluation assessments will be blinded throughout the study until the final assessment, which is performed for the control arm only. However, because the participants are not blinded, there is some risk they may reveal their intervention status to the assessors. The statisticians will be blinded during the analysis.

#### Adherence

The intervention group supervisors and research coordinator will monitor participant intervention attendance. Intervention supervisors are certified ESDM-therapists, and will follow the apprenticeship model for lay counsellor supervision in mental health, tailoring supervision intensity based on demonstrated ECD practitioner coaching competencies [58]. Adherence will be measured based on the number of coaching sessions attended, and the time period over which sessions are completed. The research coordinator will monitor attendance and completion of assessment sessions and related study protocols across participant groups. If non-adherence is observed, the research team will assess and respond at the time, and consider implementing additional outreach activities to reengage participants, but do not anticipate this to be a significant problem based on pilot work.

#### Data management

The research coordinator will oversee screening, consent, scheduling, and REDCap group allocation. The research coordinator will also oversee training of the clinical assessors and ECD practitioners. Clinical assessors, blinded to study arm allocation, will complete and rate primary and secondary clinical outcome measures. All study staff will receive training and ongoing supervision in data security and confidentiality, including confidentiality protection, mandated reporting, and ethical research conduct. All data will be coded with a unique participant ID number. The key linking names and participant IDs will be stored in REDCap and in a password protected document, accessible only to essential study staff. Surveys and assessment forms will be entered into REDCap to be stored on a private, firewall protected network. REDCap is a HIPAA compliant, web-based application.; All REDCap data transmission is encrypted, and access to the application is password-protected with a two-factor identification requirement. REDCap offers additional features for data security and management, including audit trails tracking data updates and exports, the ability to limit data access by user roles and to limit exports to deidentified data for data sharing and analysis. In accordance with terms and conditions for data sharing, other electronic data such as video recordings will be transferred to a secure, HIPAA-compliant project folder on box.com at Duke University through Box Sync, and deleted from the recording equipment post-transfer. No electronic data will be stored on a laptop or external hard drive. Duke University will manage the infrastructure where electronic research data is stored on a secure server, protected by two factor authentication. Essential study staff will access the drive through a secure VPN connection. Locked file cabinets in locked research offices will be used to securely store non-electronic data. Access to data storage areas will be restricted to essential study staff.

### Data Analyses

#### Clinical Outcomes

We will use the linear mixed-effects model framework to analyze primary and secondary child and caregiver clinical outcomes, all of which are either continuous or score outcomes. Separate models will be fitted for each outcome and will include both 16- and 24-week follow-up data collected in both trial arms, with sensitivity analyses including the additional follow-up data collected in the control arm after introduction of the intervention to that arm. The model structure will follow the approach of Moerbeek & Teernestra for an individually randomized group-treatment design with clustering in one intervention arm, by using random effects to account for the partially nested design (i.e. for ECD practitioner in the intervention arm), for correlation within-child over time, and by allowing for heteroscedasticity in residual variances in each arm [86]. Fixed effects will include intervention arm (categorical), time (categorical), the arm-by-time interaction (to allow for different intervention effects at 16- and 24-weeks), caregiver-reported child physical sex at birth (to account for the stratified design) and the baseline level of the outcome (i.e. using an analysis of covariance (ANCOVA) approach to gain power for our outcome analyses). Sensitivity analyses will additionally adjust for hours of non-study intervention services received.

Regarding missing outcome data, we note that the linear mixed effects modeling approach is valid under the missing-at-random assumption [87]. We will carefully characterize missing data patterns by arm and will summarize the baseline covariates for those with and without missing outcome data in order to identify baseline covariates that may predict missing data patterns by arm. If identified, these covariates will be additionally adjusted for in regression models. In the unlikely case that there are missing baseline levels of an outcome, the ANCOVA approach adjusting for the baseline level of the outcome would exclude all follow-up outcomes from the analysis for such participants. Were such a situation to arise, an alternative constrained longitudinal data analysis approach will be used, whereby the baseline level is modeled as an outcome with mean levels constrained to be equal in both trial arms [88].

The decision rule for our co-primary outcomes will follow the union-intersection testing approach. That is, we will consider the intervention to be successful if there is a significant signal on at-least-one of our co-primary outcomes, regardless of whether or not the other outcome is also significant. Regression diagnostics through visual inspection of the model residuals will be used to examine model assumptions, including the degree to which the fitted variance structure matches that of the data. In the case that the fit of the model is deemed to be inadequate or the model fails to converge, alternative modeling strategies will be employed (for example, generalized estimating equations). Intention-to-treat principle will be applied to all main analyses, with sensitivity analyses that adjust for compliance in the intervention arm [88].

Additional pre-planned analyses include both moderation and mediation. Potential moderators include caregiver characteristics (for example, stress, strain and sense of competence), and variability in child factors that can be measured from a dimensional perspective (for example, with the SRS-2). Each of these will be considered separately by adding the potential moderator and its interaction with the intervention indicator and with time, as well as the three-way interaction. These analyses will be completed separately for each of the two primary outcomes. A mediation analysis will be performed using 16-week uptake in caregiver strategies as a mediator on the pathway between the intervention and each of the 24-week co-primary distal outcomes. Additional exploratory analyses will include visual examination (for example, scatterplots) of changes in the SenseToKnow dimensional quantitative measures of autism-related behaviours.

#### Implementation Outcomes

Mixed methods will be used to identify implementation determinants. Qualitative data will be analyzed using directed content analysis [89]. Quantitative measures will be summarized using descriptive statistics. Three main strategies will be used to integrate qualitative and quantitative data [90]. First, through convergence, qualitative and quantitative will be triangulated to understand whether these data supply the same answer to the same question. Second, through expansion, results will be linked when the former is used to explain unexpected findings. Third, through complementarity, qualitative results will be embedded within quantitative results to contextualize quantitative results.

#### Monitoring

The study is considered minimal risk. As such, the Principal Investigators will primarily be responsible for oversight of study conduct, and will monitor the safety and data integrity for the project. The Principal Investigators will meet regularly with the study team to discuss study progress, including any adverse events or data integrity concerns. Adverse events will be monitored through a general inquiry approach, by study staff at each contact point [67]. All deviations and/or corrective actions requiring reporting will be submitted to UCT Human Research Ethics Committee (HREC) and the Duke Health System Institutional Review Board (DUHS IRB). At least twice a year, and any time an adverse event occurs, the Principal Investigators will conduct a data and safety review. During this review process, the Principal Investigators will evaluate any adverse events and determine whether these events affect the study risk/benefit ratio and whether the protocol or consent should be modified. UCT HREC and the DUHS IRB will also review this protocol on a yearly basis (or more frequently if deemed necessary) as part of the continuing review process. At this time, they will: (1) reassess the risks and benefits to participants, the informed consent process, and safeguards for human subjects, (2) review participant enrolment and retention, (3) consider any new academic developments that might impact participant safety, and (4) review any adverse events.

#### Ethics and funding

Ethics approval has been obtained from the UCT HREC (HREC REF:367/2022) and Duke IRB (Pro00111048). Additional approvals were obtained from Red Cross War Memorial Children’s Hospital (RXH: RCC358/ WC_202208_038), and the Western Cape Education Department (Reference#: 20220511-2021). The study is registered as a clinical trial on ClinicalTrials.gov (NCT05551728). Any protocol amendments will be submitted to ethics boards and the clinical trial registry. Participants will sign informed consent prior to initiation of study activities. Participants enrolled in the study will have the opportunity to evaluate and review the consent for the study, and privacy will be maintained during the conduct of intervention sessions and study related activities. For caregiver-child dyads, the child’s caregiver/legal guardian who signs the consent form must be legally authorized to sign informed consent on behalf of the child. Verbal assent from children will not be sought as per UCT Policy (study site and primary ethics board) the earliest age at which assent is recommended is 7 years, and all children in this study will be 6 years or younger at time of consent [91]. Verification of comprehension of consent will be accomplished by providing participants with a one-page summary of the informed consent information and asking participants to recall central points in the consent process. Participants will be reminded that they can stop participating at any time or refuse to attend any portion of the study or skip any question in an assessment. Any declaration of interests will be specified in consent documentation. This study has been funded by the National Institute of Health (Grant number: R01MH127573) in the United States.

## Results

In terms of current study progress: ethics approval has been obtained from the UCT and Duke, study staff have been hired, study clinicians have been trained in the administration of primary outcome measures, schools have been identified where caregiver coaching will occur, and ECD practitioners have been trained to deliver the coaching intervention. Participant enrolment was initiated in April 2023. Estimated primary completion date is March 2027. Data generated by the ACACIA trial will be shared through presentations at community events and scientific meetings, peer-reviewed publications, and will be submitted to ClinicalTrials.gov.

## Discussion

Task-sharing early autism caregiver coaching to non-specialists, integrated within existing care systems, may support scale up of services for young autistic children in low-resource environments [1, 23]. The ACACIA trial, which is adequately powered to assess the effectiveness of a contextually adapted, cascaded task-sharing intervention delivered in an educational setting, will determine whether the intervention results in significant and meaningful improvement in short-and-long-term child outcomes, and in particular meaningful improvements in child communication. Furthermore, the study will identify key determinants (barriers and facilitators) of intervention implementation, which will support the design of implementation strategies for scale up of the approach in low-resource settings.

## Limitations

Due to the nature of the coaching intervention, participants cannot be blinded. Clinical assessors are however, and we will ask caregivers not to disclose which intervention arm they are in to the assessor. Second, caregiver-child dyads may receive ongoing services (for example, speech therapy or occupational therapy) as part of usual care, which may affect child and caregiver outcomes. Moreover, the intensity, duration and focus of usual care may differ amongst participants. Third, while intervention caregivers will be asked to complete a home recorded caregiver-child interaction each week that would be reviewed in session, as an effort to support home practice and generalization of skills, they may decide not to do so, or may experience technical difficulties, thereby decreasing home-practice of strategies. Fourth, social desirability bias may impact ratings of self-reported outcomes. However, both primary outcomes are clinician administered and rated.

## Conclusions

The ACACIA trial will provide an evaluation of contextually adapted, cascaded task-sharing intervention in South Africa that can provide accessible, low-cost and scalable early autism intervention access in low-resource settings. Effective services and supports, provided in a timely way, to those in need, may improve both child and family outcomes, including quality of life.

## Authors’ contributions

**Conceptualization**: Lauren Franz, Petrus J de Vries

**Data curation**: Marisa Viljoen, Sandy Askew, Musaddiqah Brown, Elizabeth L Turner

**Funding acquisition**: Lauren Franz

**Investigation**: Marisa Viljoen, Musaddiqah Brown, Katlego Sebolai, Noleen Seris, Nokuthula Shabalala

**Methodology**: Lauren Franz, Geraldine Dawson, J Matias Di Martino, Guillermo Sapiro, Aubyn Stahmer, Elizabeth L Turner, Petrus J de Vries

**Project administration**: Lauren Franz, Marisa Viljoen, Musaddiqah Brown, Katlego Sebolai, Petrus J de Vries

**Writing – original draft:** Lauren Franz, Marisa Viljoen, Sandy Askew, Elizabeth L Turner, Petrus J de Vries

**Writing – review and editing:** Lauren Franz, Marisa Viljoen, Sandy Askew, Musaddiqah Brown, Geraldine Dawson, J Matias Di Martino, Guillermo Sapiro, Katlego Sebolai, Noleen Seris, Nokuthula Shabalala, Aubyn Stahmer, Elizabeth L Turner, Petrus J de Vries

## Supporting information

S1 Checklist. SPIRIT 2013 checklist: Recommended items to address in a clinical trial protocol and related documents

## Data Availability

Deidentified research data will be made publicly available when the study is completed and published.

## References

[1] Lord C, Charman T, Havdahl A, Carbone P, Anagnostou E, Boyd B, et al. The Lancet Commission on the future of care and clinical research in autism. Lancet. 2022 Jan 15;399(10321):271–334.

[2] Estes A, Munson J, Rogers SJ, Greenson J, Winter J, Dawson G. Long-Term Outcomes of Early Intervention in 6-Year-Old Children With Autism Spectrum Disorder. J Am Acad Child Adolesc Psychiatry. 2015 Jul;54(7):580–7.

[3] Cidav Z, Munson J, Estes A, Dawson G, Rogers S, Mandell D. Cost Offset Associated With Early Start Denver Model for Children With Autism. J Am Acad Child Adolesc Psychiatry. 2017 Sep;56(9):777–83.

[4] Franz L, Chambers N, von Isenburg M, de Vries PJ. Autism spectrum disorder in Sub-Saharan Africa: A comprehensive scoping review. Autism Res. 2017 May;10(5):723–49.

[5] United Nations Children’s Fund. UNICEF DATA. 2014 [cited 2023 Jul 3]. Generation 2030 | Africa: Child demographics in Africa. Available from: https://data.unicef.org/resources/generation-2030-africa-child-demographics-in-africa/

[6] Pacia C, Holloway J, Gunning C, Lee H. A Systematic Review of Family-Mediated Social Communication Interventions for Young Children with Autism. Rev J Autism Dev Disord. 2022;9(2):208–34.

[7] Wetherby AM, Guthrie W, Woods J, Schatschneider C, Holland RD, Morgan L, et al. Parent-implemented social intervention for toddlers with autism: an RCT. Pediatrics. 2014 Dec;134(6):1084–93.

[8] Pickles A, Le Couteur A, Leadbitter K, Salomone E, Cole-Fletcher R, Tobin H, et al. Parent-mediated social communication therapy for young children with autism (PACT): long-term follow-up of a randomised controlled trial. Lancet. 2016 Nov 19;388(10059):2501–9.

[9] Schreibman L, Dawson G, Stahmer AC, Landa R, Rogers SJ, McGee GG, et al. Naturalistic Developmental Behavioral Interventions: Empirically Validated Treatments for Autism Spectrum Disorder. J Autism Dev Disord. 2015;45(8):2411–28.

[10] Dawson G, Rogers S, Munson J, Smith M, Winter J, Greenson J, et al. Randomized, controlled trial of an intervention for toddlers with autism: the Early Start Denver Model. Pediatrics. 2010 Jan;125(1):e17–23.

[11] Shi B, Wu W, Dai M, Zeng J, Luo J, Cai L, et al. Cognitive, Language, and Behavioral Outcomes in Children With Autism Spectrum Disorders Exposed to Early Comprehensive Treatment Models: A Meta-Analysis and Meta-Regression. Front Psychiatry. 2021;12:691148.

[12] Rogers SJ, Estes A, Lord C, Munson J, Rocha M, Winter J, et al. A Multisite Randomized Controlled Two-Phase Trial of the Early Start Denver Model Compared to Treatment as Usual. J Am Acad Child Adolesc Psychiatry. 2019 Sep;58(9):853–65.

[13] Sinai-Gavrilov Y, Gev T, Mor-Snir I, Vivanti G, Golan O. Integrating the Early Start Denver Model into Israeli community autism spectrum disorder preschools: Effectiveness and treatment response predictors. Autism. 2020 Nov;24(8):2081–93.

[14] Lin TL, Chiang CH, Ho SY, Wu HC, Wong CC. Preliminary clinical outcomes of a short-term low-intensity Early Start Denver Model implemented in the Taiwanese public health system. Autism. 2020 Jul;24(5):1300–6.

[15] Colombi C, Narzisi A, Ruta L, Cigala V, Gagliano A, Pioggia G, et al. Implementation of the Early Start Denver Model in an Italian community. Autism. 2018 Feb;22(2):126–33.

[16] Devescovi R, Colonna V, Dissegna A, Bresciani G, Carrozzi M, Colombi C. Feasibility and Outcomes of the Early Start Denver Model Delivered within the Public Health System of the Friuli Venezia Giulia Italian Region. Brain Sci. 2021 Sep 10;11(9):1191.

[17] Zhou B, Xu Q, Li H, Zhang Y, Wang Y, Rogers SJ, et al. Effects of Parent-Implemented Early Start Denver Model Intervention on Chinese Toddlers with Autism Spectrum Disorder: A Non-Randomized Controlled Trial: Effects of P-ESDM in China. Autism Research. 2018 Apr;11(4):654–66.

[18] Tateno Y, Kumagai K, Monden R, Nanba K, Yano A, Shiraishi E, et al. The Efficacy of Early Start Denver Model Intervention in Young Children with Autism Spectrum Disorder Within Japan: A Preliminary Study. Soa Chongsonyon Chongsin Uihak. 2021 Jan 1;32(1):35–40.

[19] Holzinger D, Laister D, Vivanti G, Barbaresi WJ, Fellinger J. Feasibility and Outcomes of the Early Start Denver Model Implemented with Low Intensity in a Community Setting in Austria. J Dev Behav Pediatr. 2019 Jun;40(5):354–63.

[20] Vivanti G, Dissanayake C, Duncan E, Feary J, Capes K, Upson S, et al. Outcomes of children receiving Group-Early Start Denver Model in an inclusive versus autism-specific setting: A pilot randomized controlled trial. Autism. 2019 Jul;23(5):1165–75.

[21] Roche L, Adams D, Clark M. Research priorities of the autism community: A systematic review of key stakeholder perspectives. Autism. 2021 Feb;25(2):336–48.

[22] Schuck RK, Tagavi DM, Baiden KMP, Dwyer P, Williams ZJ, Osuna A, et al. Neurodiversity and Autism Intervention: Reconciling Perspectives Through a Naturalistic Developmental Behavioral Intervention Framework. J Autism Dev Disord. 2022 Oct;52(10):4625–45.

[23] Rieder AD, Viljoen M, Seris N, Shabalala N, Ndlovu M, Turner EL, et al. Improving access to early intervention for autism: findings from a proof-of-principle cascaded task-sharing naturalistic developmental behavioural intervention in South Africa. Child Adolesc Psychiatry Ment Health. 2023 May 20;17(1):64.

[24] Reichow B, Servili C, Yasamy MT, Barbui C, Saxena S. Non-specialist psychosocial interventions for children and adolescents with intellectual disability or lower-functioning autism spectrum disorders: a systematic review. PLoS Med. 2013 Dec;10(12):e1001572; discussion e1001572.

[25] Wong VCN, Kwan QK. Randomized controlled trial for early intervention for autism: a pilot study of the Autism 1-2-3 Project. J Autism Dev Disord. 2010 Jun;40(6):677–88.

[26] Tsang SKM, Shek DTL, Lam LL, Tang FLY, Cheung PMP. Brief Report: Application of the TEACCH Program on Chinese Pre-School Children with Autism––Does Culture Make a Difference? J Autism Dev Disord. 2007 Feb 1;37(2):390–6.

[27] Divan G, Vajaratkar V, Cardozo P, Huzurbazar S, Verma M, Howarth E, et al. The Feasibility and Effectiveness of PASS Plus, A Lay Health Worker Delivered Comprehensive Intervention for Autism Spectrum Disorders: Pilot RCT in a Rural Low and Middle Income Country Setting. Autism Res. 2019 Feb;12(2):328–39.

[28] Schleiff MJ, Aitken I, Alam MA, Damtew ZA, Perry HB. Community health workers at the dawn of a new era: 6. Recruitment, training, and continuing education. Health Research Policy and Systems. 2021 Oct 12;19(3):113.

[29] Black MM, Walker SP, Fernald LCH, Andersen CT, DiGirolamo AM, Lu C, et al. Early childhood development coming of age: science through the life course. Lancet. 2017 Jan 7;389(10064):77–90.

[30] Richter LM, Daelmans B, Lombardi J, Heymann J, Boo FL, Behrman JR, et al. Investing in the foundation of sustainable development: pathways to scale up for early childhood development. Lancet. 2017 Jan 7;389(10064):103–18.

[31] Jaskiewicz W, Tulenko K. Increasing community health worker productivity and effectiveness: a review of the influence of the work environment. Human Resources for Health. 2012 Sep 27;10(1):38.

[32] Smith S, Deveridge A, Berman J, Negin J, Mwambene N, Chingaipe E, et al. Task-shifting and prioritization: a situational analysis examining the role and experiences of community health workers in Malawi. Human Resources for Health. 2014 May 2;12(1):24.

[33] Okyere E, Mwanri L, Ward P. Is task-shifting a solution to the health workers’ shortage in Northern Ghana? PLoS One. 2017 Mar 30;12(3):e0174631.

[34] The World Bank. World Development Indicators | DataBank [Internet]. [cited 2023 Jul 3]. Available from: https://databank.worldbank.org/source/world-development-indicators.%20Accessed%209December%202022

[35] Guler J, de Vries PJ, Seris N, Shabalala N, Franz L. The importance of context in early autism intervention: A qualitative South African study. Autism. 2018 Nov;22(8):1005–17.

[36] Franz L, Adewumi K, Chambers N, Viljoen M, Baumgartner JN, de Vries PJ. Providing early detection and early intervention for autism spectrum disorder in South Africa: stakeholder perspectives from the Western Cape province. J Child Adolesc Ment Health. 2018 Nov;30(3):149–65.

[37] Pillay S, Duncan M, de Vries PJ. Autism in the Western Cape province of South Africa: Rates, socio-demographics, disability and educational characteristics in one million school children. Autism. 2021 May;25(4):1076–89.

[38] Pillay S, Duncan M, de Vries PJ. Who’s waiting for a school? Rates, socio-demographics, disability and referral profile of children with autism spectrum disorder awaiting school placement in the Western Cape Province of South Africa. Autism. 2022 Oct;26(7):1849–63.

[39] Pillay S, Duncan M, de Vries PJ. “We are doing the best we can to bridge the gap” - service provider perspectives of educational services for autism spectrum disorder in South Africa. Front Psychiatry. 2022;13:907093.

[40] Mayosi BM, Benatar SR. Health and Health Care in South Africa — 20 Years after Mandela. New England Journal of Medicine. 2014 Oct 2;371(14):1344–53.

[41] Springer PE, van Toorn R, Laughton B, Kidd M. Characteristics of children with pervasive developmental disorders attending a developmental clinic in the Western Cape Province, South Africa. South African Journal of Child Health. 2013 Aug 30;7(3):95–9.

[42] Guler J, Stewart KA, de Vries PJ, Seris N, Shabalala N, Franz L. Conducting caregiver focus groups on autism in the context of an international research collaboration: Logistical and methodological lessons learned in South Africa. Autism. 2023 Apr;27(3):751–61.

[43] Department of Social Development. National Integrated Early Childhood Development Policy | UNICEF South Africa [Internet]. [cited 2023 Jul 3]. Available from: https://www.unicef.org/southafrica/reports/national-integrated-early-childhood-development-policy

[44] South African Government. Joint Press Statement: Early Childhood Development (ECD) Function Shift | South African Government [Internet]. [cited 2023 Jul 3]. Available from: https://www.gov.za/speeches/joint-press-statement-early-childhood-development-ecd-function-shift-16-mar-2021-0000

[45] Berg A, Lachman A. Positive Relational Experiences in Infancy May Influence Outcomes in Children in a Low and Middle-Income Country Setting Such as South Africa. Front Public Health. 2021;9:665908.

[46] Ramseur K, de Vries PJ, Guler J, Shabalala N, Seris N, Franz L. Caregiver descriptions of joint activity routines with young children with autism spectrum disorder in South Africa. Pediatr Med. 2019 Mar;2:6.

[47] Makombe CBT, Shabalala N, Viljoen M, Seris N, de Vries PJ, Franz L. Sustainable implementation of early intervention for autism spectrum disorder through caregiver coaching: South African perspectives on barriers and facilitators. Pediatr Med. 2019 Aug;2:39.

[48] Ndlovu M. Towards Naturalistic Developmental Behavioural Interventions for Autism in Africa: Nature and Context of Caregiver-Child Interactions in Low-Resource South African Environments [MSc (Med) Neuroscience]. [Cape Town, South Africa]: University of Cape Town; 2022.

[49] Sparrow SS, Cichetti S, Saulnier C. Vineland Adaptive Behavior Scales, Third Edition. San Antonio, TX: Pearson.

[50] Green E, Stroud L, Bloomfield S, Cronje J, Foxcroft C, Hurter K, et al. Griffiths Scales of Child Development, Third Edition. Göttingen: Hogrefe Publishing Group.

[51] Franz L, Goodwin CD, Rieder A, Matheis M, Damiano DL. Early intervention for very young children with or at high likelihood for autism spectrum disorder: An overview of reviews. Dev Med Child Neurol. 2022 Sep;64(9):1063–76.

[52] Sandbank M, Bottema-Beutel K, Crowley S, Cassidy M, Dunham K, Feldman JI, et al. Project AIM: Autism intervention meta-analysis for studies of young children. Psychol Bull. 2020 Jan;146(1):1–29.

[53] Aarons GA, Hurlburt M, Horwitz SM. Advancing a conceptual model of evidence-based practice implementation in public service sectors. Adm Policy Ment Health. 2011 Jan;38(1):4–23.

[54] Moullin JC, Dickson KS, Stadnick NA, Rabin B, Aarons GA. Systematic review of the Exploration, Preparation, Implementation, Sustainment (EPIS) framework. Implement Sci. 2019 Jan 5;14(1):1.

[55] Dingfelder HE, Mandell DS. Bridging the Research-to-Practice Gap in Autism Intervention: An Application of Diffusion of Innovation Theory. J Autism Dev Disord. 2011 May;41(5):597–609.

[56] Curran GM, Bauer M, Mittman B, Pyne JM, Stetler C. Effectiveness-implementation hybrid designs: combining elements of clinical effectiveness and implementation research to enhance public health impact. Med Care. 2012 Mar;50(3):217–26.

[57] Pals SL, Murray DM, Alfano CM, Shadish WR, Hannan PJ, Baker WL. Individually Randomized Group Treatment Trials: A Critical Appraisal of Frequently Used Design and Analytic Approaches. Am J Public Health. 2008 Aug;98(8):1418–24.

[58] Murray LK, Dorsey S, Bolton P, Jordans MJ, Rahman A, Bass J, et al. Building capacity in mental health interventions in low resource countries: an apprenticeship model for training local providers. International Journal of Mental Health Systems. 2011 Nov 18;5(1):30.

[59] Adamson LB, Bakeman R, Deckner DF, Nelson PB. Rating parent-child interactions: joint engagement, communication dynamics, and shared topics in autism, Down syndrome, and typical development. J Autism Dev Disord. 2012 Dec;42(12):2622–35.

[60] Chan AW, Tetzlaff JM, Altman DG, Laupacis A, Gøtzsche PC, Krleža-Jerić K, et al. SPIRIT 2013 Statement: Defining Standard Protocol Items for Clinical Trials. Ann Intern Med. 2013 Feb 5;158(3):200–7.

[61] American Psychiatric Association. Diagnostic and Statistical Manual of Mental Disorders (DSM-5). Arlington, VA: American Psychiatric Association;

[62] Lord C, Rutter M, DiLavore P, Risi S, Gotham K, Bishop S. Autism diagnostic observation schedule, Second Edition. Torrance, CA: Western Psychological Services; 2012.

[63] Rogers S, Stahmer A. Help Is In Your Hands | Login [Internet]. [cited 2023 Jul 19]. Available from: https://helpisinyourhands.org/course

[64] Rogers SJ, Stahmer A, Talbott M, Young G, Fuller E, Pellecchia M, et al. Feasibility of delivering parent-implemented NDBI interventions in low-resource regions: a pilot randomized controlled study. J Neurodev Disord. 2022 Jan 5;14(1):3.

[65] Frost KM, Brian J, Gengoux GW, Hardan A, Rieth SR, Stahmer A, et al. Identifying and measuring the common elements of naturalistic developmental behavioral interventions for autism spectrum disorder: Development of the NDBI-Fi. Autism. 2020 Nov;24(8):2285–97.

[66] Bernal G, Bonilla J, Bellido C. Ecological validity and cultural sensitivity for outcome research: issues for the cultural adaptation and development of psychosocial treatments with Hispanics. J Abnorm Child Psychol. 1995 Feb;23(1):67–82.

[67] Coates M, Spanos M, Parmar P, Chandrasekhar T, Sikich L. A Review of Methods for Monitoring Adverse Events in Pediatric Psychopharmacology Clinical Trials. Drug Saf. 2018;41(5):465–71.

[68] Johnston C, Mash EJ. A Measure of Parenting Satisfaction and Efficacy. Journal of Clinical Child Psychology. 1989 Jun 1;18(2):167–75.

[69] Abidin R. Manual for the parenting stress index. 3rd ed. Odessa, FL: Psychological Assessment Resources; 1995.

[70] Brannan AM, Heflinger CA, Brickman L. The Caregiver Strain Questionnaire: Measuring the Impact on the Family of Living with a Child with Serious Emotional Disturbance. Journal of Emotional and Behavioral Disorders. 1997;5(4):212–22.

[71] Constantino J, Gruber C. Social responsiveness scale (SRS) manual. Los Angeles, CA: Western Psychological Services; 2005.

[72] Perochon S, Di Martino M, Aiello R, Baker J, Carpenter K, Chang Z, et al. A scalable computational approach to assessing response to name in toddlers with autism. J Child Psychol Psychiatry. 2021 Sep;62(9):1120–31.

[73] Perochon S, Matias Di Martino J, Carpenter KLH, Compton S, Davis N, Espinosa S, et al. A tablet-based game for the assessment of visual motor skills in autistic children. NPJ Digit Med. 2023 Feb 3;6(1):17.

[74] Krishnappa Babu PR, Di Martino JM, Chang Z, Perochon S, Aiello R, Carpenter KLH, et al. Complexity analysis of head movements in autistic toddlers. J Child Psychol Psychiatry. 2023 Jan;64(1):156–66.

[75] Chang Z, Di Martino JM, Aiello R, Baker J, Carpenter K, Compton S, et al. Computational Methods to Measure Patterns of Gaze in Toddlers With Autism Spectrum Disorder. JAMA Pediatr. 2021 Aug 1;175(8):827–36.

[76] Carpenter KLH, Hahemi J, Campbell K, Lippmann SJ, Baker JP, Egger HL, et al. Digital Behavioral Phenotyping Detects Atypical Pattern of Facial Expression in Toddlers with Autism. Autism Res. 2021 Mar;14(3):488– 99.

[77] Perochon S, Di Martino JM, Carpenter KLH, Compton S, Davis N, Eichner B, et al. Early detection of autism using digital behavioral phenotyping. Nature Medicine, In Press.

[78] Aarons GA, Ehrhart MG, Farahnak LR. The implementation leadership scale (ILS): development of a brief measure of unit level implementation leadership. Implementation Science. 2014 Apr 14;9(1):45.

[79] Ehrhart MG, Aarons GA, Farahnak LR. Assessing the organizational context for EBP implementation: the development and validity testing of the Implementation Climate Scale (ICS). Implementation Science. 2014 Oct 23;9(1):157.

[80] Rogers SJ, Vismara LA, Dawson G. Coaching parents of young children with autism: promoting connection, communication, and learning. New York: Guilford Publications; 2021.

[81] Drahota A, Meza RD, Brikho B, Naaf M, Estabillo JA, Gomez ED, et al. Community-Academic Partnerships: A Systematic Review of the State of the Literature and Recommendations for Future Research. Milbank Q. 2016 Mar;94(1):163–214.

[82] Gomez E, Drahota A, Stahmer AC. Choosing strategies that work from the start: A mixed methods study to understand effective development of community-academic partnerships. Action Res (Lond). 2021 Jun;19(2):277– 300.

[83] National Institutes of Health. IRGT Sample Size Calculator, Research Methods Resources [Internet]. 2022 [cited 2023 Aug 17]. Available from: https://researchmethodsresources.nih.gov/Tools#irgt

[84] Harris PA, Taylor R, Thielke R, Payne J, Gonzalez N, Conde JG. Research electronic data capture (REDCap)--a metadata-driven methodology and workflow process for providing translational research informatics support. J Biomed Inform. 2009 Apr;42(2):377–81.

[85] Harris PA, Taylor R, Minor BL, Elliott V, Fernandez M, O’Neal L, et al. The REDCap consortium: Building an international community of software platform partners. J Biomed Inform. 2019 Jul;95:103208.

[86] Moerbeek M, Teerenstra S. Power analysis of tials with multilevel data. Boca Raton, FL: Chapman & Hall/CRC Press; 2016.

[87] Fitzmaurice G, Laird N, Ware J. Applied Longitudinal Analysis, 2nd Edition. Wiley; 2011.

[88] Coffman CJ, Edelman D, Woolson RF. To condition or not condition? Analysing ‘change’ in longitudinal randomised controlled trials. BMJ Open. 2016 Dec 1;6(12):e013096.

[89] Hsieh HF, Shannon SE. Three approaches to qualitative content analysis. Qual Health Res. 2005 Nov;15(9):1277–88.

[90] Creswell J, Plano Clark V. Designing and conducting mixed methods research (2nd edition). Los Angeles, CA: SAGE Publications; 2011.

[91] University of Cape Town, Faculty of Health Sciences. Human Research Ethics Committee Standard Operating Procedures Version 7.0 [Internet]. 2019. Available from: https://health.uct.ac.za/sites/default/files/media/documents/health_uct_ac_za/54/UCT%20HREC%20Standard%20Operating%20Procedures%20Version%207.0%20October%202019.pdf

